# Effects of spinal stimulation and short-burst treadmill training on gait biomechanics in children with cerebral palsy

**DOI:** 10.1101/2024.05.31.24308292

**Authors:** Charlotte R. DeVol, Siddhi R. Shrivastav, Victoria M. Landrum, Kristie F. Bjornson, Desiree Roge, Chet T. Moritz, Katherine M. Steele

## Abstract

**Background:** Children with cerebral palsy (CP) have an injury to the central nervous system around the time of birth that affects the development of the brain and spinal cord. This injury leads to changes in gait neuromechanics, including muscle activity and joint kinematics. Transcutaneous spinal cord stimulation (tSCS) is a novel neuromodulation technique that may improve movement and coordination in children with CP when paired with targeted physical therapy.

**Research question:** How does the combination of tSCS and short-burst interval locomotor treadmill training (SBLTT) affect individual gait neuromechanics in children with CP?

**Methods:** Four children with CP (4-13 years old), received 24 sessions each of SBLTT only and SBLTT with tSCS (tSCS+SBLTT). Clinical assessments of spasticity and passive range of motion (PROM), as well as biomechanical assessments of joint kinematics, musculotendon lengths, and muscle activity were recorded during overground, barefoot walking. Assessments were taken before and after each intervention, and 8-weeks later.

**Results:** The combination of tSCS+SBLTT led to greater increases in hip and knee extension than SBLTT only for three participants. Three children also became more plantarflexed at the ankle during stance after tSCS+SBLTT compared to SBLTT only. While tSCS+SBLTT reduced spasticity, these changes were only weakly correlated with changes in musculotendon lengths during gait or PROM, with the largest correlation between change in gastrocnemius operating musculotendon length during fast walking and gastrocnemius spasticity (R^2^ = 0.26) and change in plantarflexor PROM and gastrocnemius spasticity (R^2^ = 0.23).

**Significance:** Children with CP used a more upright, less crouched posture during gait after tSCS+SBLTT. Large reductions in spasticity after tSCS+SBLTT were only weakly correlated with changes in kinematics and PROM. Understanding the mechanisms by which tSCS may affect gait for children with CP is critical to optimize and inform the use of tSCS for clinical care.

## Introduction

Cerebral Palsy (CP) is caused by an injury to the central nervous system around the time of birth that affects the control of movement. In spastic CP, this initial injury leads to neuromuscular impairments of weakness, shortened musculotendon units, spasticity, and impaired selective motor control, as well as secondary effects of joint contracture and bone deformity that contribute to altered gait mechanics and coordination causing fatiguing gait patterns in children with CP [1–4].

Transcutaneous spinal cord stimulation (tSCS) is a novel intervention that may support reorganization of neural pathways to improve movement coordination for children with CP [5–7]. A single session of tSCS has been shown to improve coordination between the hip and knee during gait and reduce inefficient muscle co-contraction [5]. Prior work hypothesized that tSCS supports activity-dependent neuroplasticity by inducing reorganization of the central nervous system through increased sensory feedback [8]. Repeated delivery of the combination of tSCS and physical therapy has led to improvements in gross motor function for children with CP [6,7,9]. Understanding the neuromechanical response to therapeutic tSCS is critical in defining the mechanisms by which spinal stimulation may improve gross motor function.

Our prior work has shown preliminary evidence that combining tSCS with treadmill training can reduce spasticity while maintaining clinical measures of walking function [10]. We evaluated the combined effects of tSCS with short-burst interval locomotor treadmill training (SBLTT). SBLTT is designed to mimic natural variability of a child’s walking pattern, providing intensive walking practice alternating between slow and fast speeds [11]. Prior work has demonstrated how evaluating changes in gait neuromechanics – like joint kinematics, muscle activity, or musculotendon lengths (MTLs) can provide key insights into the mechanisms by which interventions like orthopedic surgery [12,13] or spasticity treatments [14] impact gait and inform clinical decision-making [15,16]. Reductions in spasticity, such as those reported after botulinum toxin type-A injections (BoNT-A) or selective dorsal rhizotomy (SDR), may increase passive range of motion (PROM) or increase joint extension and musculotendon operating lengths (MTLs) during gait [17–21].

The purpose of this pilot study was to quantify the effects of SBLTT with and without tSCS on gait kinematics, muscle activity, MTLs, and PROM in children with CP and how these changes relate to changes in spasticity. We hypothesized that reductions in spasticity with tSCS+SBLTT would be associated with increases in PROM, greater extension in the hip and knee throughout the gait cycle, reductions in ankle dorsiflexion during stance, and increases in MTLs during gait.

## Methods

### Study Design

This study quantifies gait biomechanics collected during a preliminary investigation [10] of the impacts of SBLTT first and then the additive effect of tSCS+SBLTT on spasticity and walking. Each participant received 24 sessions of SBLTT first, and then 24 sessions of tSCS+SBLTT, with an 8-week follow-up after each intervention (Figure 1A). In this small sample size, the order of treatment was kept consistent to avoid confounding carryover effects of tSCS that are known to persist for several months in people with spinal cord injury [22–24]. Clinical assessments for PROM and spasticity, as well as gait analysis of kinematics, MTLs, and muscle activity were recorded the week immediately before and after each intervention and at 8-weeks follow-up. All assessment visits were conducted at the University of Washington. This study was approved by the University of Washington Human Subjects Division (IRB identifier: STUDY00008896) and was registered on ClinicalTrials.gov (NCT04467437).

**Figure 1.**
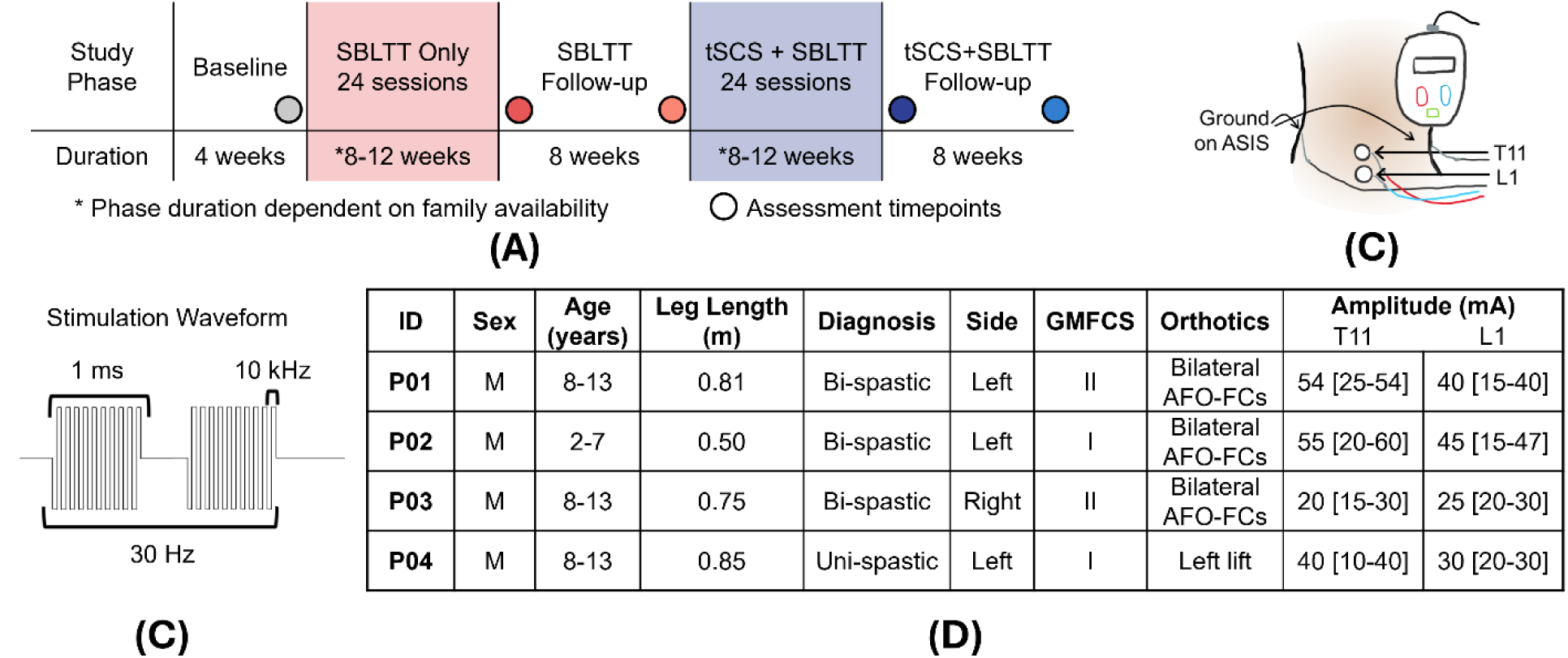
A) Study timeline indicating assessment timepoints before and after each intervention and at 8-weeks follow-up. SBLTT = short-burst interval locomotor treadmill training; tSCS = transcutaneous spinal cord stimulation. B) Diagram of investigative spinal cord neuromodulation device (SpineX, Inc.) with stimulating electrodes on the T11 and L1 dorsal spinous processes and two ground electrodes on the anterior superior iliac spine (ASIS). C) Diagram of the stimulation waveform. D) Participant information: ID = participant identifier; Bi-spastic = bilateral spastic CP; Uni-spastic = unilateral spastic CP; Side = more-affected side based on the side with more spasticity at baseline and parent reports; GMFCS = Gross Motor Function Classification System Level; The tSCS amplitude applied to T11 = thoracic spinous process 11, L1 = lumbar spinous process with values reported as median [range].

Each training session included a 5 to 15-minute active warm-up either overground or on the treadmill, 30-minutes of SBLTT, and a 5-minute active cool-down. Rest breaks were provided as needed, and participants were encouraged to use handrails for safety. SBLTT was individualized based on baseline overground walking speeds and consisted of 30-second bursts, alternating between walking at a slow and fast pace [25]. The slow pace remained constant between visits, while the fast speed increased based on perceived exertion. During training, all participants wore devices they used during community walking, including ankle foot orthoses footwear combinations and shoe lifts (Figure 1D).

During tSCS+SBLTT, the same protocol for SBLTT was followed with the addition of transcutaneous spinal cord stimulation, administered via an investigative spinal cord neuromodulation device (SpineX, Inc.) [5]. Stimulation was applied using adhesive gel electrodes placed just below the T11 and L1 spinous processes using 3.2 cm round electrodes. The ground electrodes were 5.1 x 8.6 cm rectangular electrodes placed over the anterior superior iliac spine (Axelgaard Manufacturing Co., Ltd., USA) (Figure 1B). Stimulation was delivered with a biphasic waveform at 30 Hz with a 1 ms pulse width. Each burst had a high carrier frequency at 10 kHz [5] (Figure 1C). During each visit, stimulation was applied for an average of 56 ± 10 minutes, including all training and rest breaks. Amplitude for the subthreshold stimulation was determined based on children’s self-report of quality of walking, sensation beneath the cathodes, and a physical therapist’s clinical observation of gait quality and participant’s behavior (Figure 1D).

### Participants

We enrolled four ambulatory children with spastic CP Gross Motor Function Classification System (GMFCS) Levels I-II [1] who were not regularly taking spasticity medications, did not have history of SDR, and had not undergone a lower extremity surgery or BoNT-A injections in the past year (Figure 1D). Two participants, P02 and P03, weaned off their daily use of baclofen three-weeks before starting the study. Another participant, P01 took baclofen as needed prior to the study and took 5 mg once during the SBLTT phase. P04 did not take baclofen prior to or during the study. Children and parents were informed of the study procedures and signed an informed consent and age-appropriate assent form.

### Clinical Assessments

Modified Ashworth Scale (MAS) was used to assess spasticity of the hamstrings, quadriceps, gastrocnemius, and soleus muscles. MAS scores were converted to an ordinal scale such that a value of zero indicates no spasticity and a value of five indicates a rigid joint. Passive range of motion (PROM) in hip flexion, knee flexion and extension, and ankle dorsiflexion and plantarflexion were measured while lying supine [26]. The hip was flexed when measurements were taken at the knee, and the midfoot and subtalar joints maintained in a neutral position during ankle movement.

### Gait Assessment

Sagittal plane joint kinematics and muscle activity were quantified during gait across a 10-meter walkway. Participants walked barefoot at a self-selected pace for a minimum of ten steps with the more-affected leg. Lower extremity position data were collected using a modified Helen-Hayes marker set [23] and a 10- or 12-camera motion capture system at 120 Hz (Qualisys AB, Gothenburg, SE). Electromyography (EMG) data (Delsys Inc, Natick, MA) were synchronously recorded at 2000 Hz during motion capture trials bilaterally for five muscles: rectus femoris (RF), vastus medialis (VM), biceps femoris (BF), tibialis anterior (TA), and medial gastrocnemius (MG). This was repeated at a fast-walking pace for the calculation of MTLs and musculotendon lengthening rates (MTL rates) for all participants, except P01 who did not complete fast walking trials at Baseline. Faster walking speeds can help identify functional limitations, such as whether or not spasticity is impacting MTLs and MTL rate [27,28].

### Data Analysis

Position data were processed using custom MATLAB scripts (MathWorks, Natick, MA), USA) and OpenSim v4.3 (Stanford, USA) using a 23 degree-of-freedom model scaled to each individual participant [29,30]. Across trials, the root-mean-square (RMS) and maximum model error for all markers were below 2 cm and 4 cm, respectively [31]. After inverse kinematics were calculated in OpenSim, joint kinematics and MTLs were segmented by gait cycle. The change in joint kinematics during each intervention were calculated as the difference of the average kinematic trajectories between the post-SBLTT and Baseline timepoints and the post-tSCS+SBLTT and SBLTT follow-up timepoints. The derivative of MTL with respect to time was used to calculate the musculotendon lengthening rates (MTL rates). All MTLs and MTL rates were normalized to the muscle length, *l*_*ref*_ and 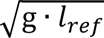, respectively, where *l*_*ref*_ is the muscle length when the participant stood in an upright posture [27]. Walking speed was calculated from the distance and time traveled of a reflective marker on the left anterior superior iliac spine for each trial and normalized to participant leg lengths as the nondimensional Froude Number [32].

Raw EMG signals were high pass filtered (4th order Butterworth; 20 Hz), zero-centered, rectified, and low pass-filtered (4th order Butterworth; 10 Hz) using custom MATLAB scripts, Signals were then normalized to the 95th percentile of maximum activation across self-selected walking speed trials for that day and reported as millivolts/millivolts (mV/mV). EMG data were segmented by gait cycle and any steps with an average standard deviation greater than one or with any single data point that exceeded four standard deviations from the mean were removed. Co-contraction of antagonistic muscle pairs was defined as the co-contraction index (CCI) calculated as:

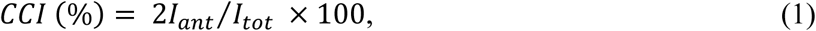

where *I_ant_* is the antagonistic muscle activity and *I_tot_* is the sum of agonist and antagonist EMG activity [33]. Reductions in CCI can suggest reduced muscle activity and improved neuromuscular control that can support a more-efficient gait pattern [34]. We also used weighted nonnegative matrix factorization with the Matrix Factorization Toolbox in MATLAB [35] to calculate muscle synergies. The total variance accounted for by one synergy (tVAF_1_) is reported as a biomarker for motor control in CP [36], with higher tVAF_1_ values suggesting less refined motor control.

### Statistical Analysis

One-dimensional statistical parametric mapping (SPM) was used to assess the difference between study time points for the kinematic trajectories over gait cycles for each individual (www.spdm1d.org in MATLAB). A normality test (spm1d.stats.normality.anova1) was used to determine whether a one-way Analysis of Variance (ANOVA) was performed or if the non-parametric version was used. If significant, post-hoc SPM two-tailed paired t-test, or the nonparametric equivalent, were used to compare changes during each intervention and follow-up relative to the pre-intervention timepoint with Bonferroni corrections for the four tests (alpha = 0.05). We used linear regression to evaluate the relationship between change in PROM, MTL, and MTL rates with spasticity. Each participant was represented in these models with two data points: one from change after SBLTT only and one from the change after tSCS+SBLTT.

## Results

Spasticity, as measured by the MAS, was reduced to a greater extent after tSCS+SBLTT compared to SBLTT only. In addition, lower spasticity was sustained for 8 weeks after tSCS+SBLTT, but not after SBLTT only (Table 1) [10]. Individual participant values in Table 1 can be found in Supplemental Tables 1-4.

**Table 1.** Average [Range] spasticity, PROM, and gait measures at each time point.

|  |  | **Baseline** | **Post-SBLTT** | **SBLTT Follow-up** | **Post-tSCS+SBLTT** | **tSCS+SBLTT Follow-up** |
| --- | --- | --- | --- | --- | --- | --- |
| **MAS** | Quadriceps | 1.25 [0-3] | 0.50 [0-2] | 0.75 [0-2] | 0 [0-0] | 0 [0-0] |
|  | Hamstrings | 1.75 [1-3] | 1.75 [0-1] | 1.25 [1-2] | 0 [0-0] | 0.75 [0-1] |
|  | Gastrocnemius | 3.50 [3-4] | 3.00 [2-4] | 3.50 [3-4] | 1.25 [1-2] | 1.25 [1-2] |
|  | Soleus | 3.50 [3-4] | 3.25 [3-4] | 3.50 [3-4] | 1.50 [1-2] | 2.00 [1-3] |
| **PROM (degrees)** | Hip Flexion | 36 [30-42] | 44 [42-45] | 40 [30-48] | 50 [42-64] | 39 [35-42] |
|  | Knee Extension | 143 [104-165] | 155 [154-155] | 149 [127-166] | 166 [155-174] | 162 [152-167] |
|  | Knee Flexion | 132 [121-153] | 129 [125-135] | 123 [110-133] | 138 [130-150] | 130 [125-140] |
|  | Ankle Dorsiflexion | 66 [50-75] | 70 [55-77] | 66 [60-80] | 84 [73-95] | 76 [65-85] |
|  | Ankle Plantarflexion | 143 [120-155] | 155 [140-268] | 149 [130-162] | 156 [152-160] | 151 [136-174] |
| **Gait Speed (n.d.)** | Self-selected Pace | 0.45 [0.31-0.52] | 0.47 [0.30-0.61] | 0.47 [0.36-0.59] | 0.49 [0.41-0.58] | 0.49 [0.39-0.61] |
|  | Fast Pace | *0.63 [0.53-0.76] | 0.64 [0.50-0.91] | 0.67 [0.54-0.84] | 0.65 [0.44-0.89] | 0.71 [0.51-0.93] |
| **Joint Kinematics (degrees)** | Peak Hip Extension | 4 [-5-16] | 4 [-8-17] | 4 [0-16] | 10 [3-15] | 12 [7-19] |
|  | Minimum Knee Flexion | 22 [18-25] | 20 [13-29] | 24 [19-30] | 16 [7-24] | 14 [9-21] |
| **Integrated EMG During Stance [% of gait cycle used]** | RF [0-60] | 24 [17-32] | 22 [18-26] | 23 [15-33] | 22 [16-33] | 23 [17-33] |
|  | VM [0-60] | *16 [11-25] | 23 [8-43] | 18 [12-25] | 15 [7-21] | 18 [10-26] |
|  | BF [0-60] | 20 [16-23] | 25 [10-37] | 21 [13-29] | 22 [15-27] | 23 [15-27] |
|  | MG [30-50] | 13 [8-15] | 12 [10-14] | 13 [10-14] | 12 [9-14] | 12 [9-15] |
|  | TA [6-60] | 23 [10-37] | 20 [12-30] | 24 [17-29] | 21 [10-35] | 21 [10-36] |
| **CCI (%)** | Proximal Muscles | *61 [52-68] | 67 [53-86] | 73 [63-81] | 55 [37-81] | 58 [36-75] |
|  | Distal Muscles | 57 [34-77] | 57 [44-74] | 67 [62-74] | 60 [43-79] | 63 [52-85] |
| **Motor Control** | tVAF<sub>1</sub> | *0.77 [0.72-0.87] | 0.82 [0.71-0.93] | 0.82 [0.74-0.92] | 0.82 [0.73-0.92] | 0.82 [0.77-0.89] |

MAS: Modified Ashworth Scale; PROM: passive range of motion with measurements at the knee taken while the hip is flexed; n.d.: nondimensionalized; EMG = electromyography; RF = rectus femoris; VM = vastus medialis; BF = bicep femoris; MG = medial gastrocnemius; TA = tibialis anterior; CCI: Co-contraction Index; Motor Control as measured by the total variance accounted for in one muscle synergy (tVAF<sub>1</sub>) on participants' more-affected side, dimensionless gait speed normalized to leg length, and peak extension during gait. \*P01 is not included in the baseline fast walking speed, and his VM EMG at Baseline had to be dropped from the EMG analysis of integrated area, proximal muscle CCI, and tVAF<sub>1</sub>.

The PROM across all joints was greater after tSCS+SBLTT compared to SBLTT only by an average of 16° for hip flexion, 11° for knee extension, 9° for knee flexion, 14° for ankle dorsiflexion, and 1° for ankle plantarflexion (Table 1). Hip flexion, knee extension, and ankle plantarflexion PROM remained greater at 8 weeks after SBLTT (average > 4°), while PROM at all joints except hip flexion remained greater at 8 weeks after tSCS+SBLTT (average > 3°). Correlations between changes in PROM and MAS were weak (Figure 4). The largest correlation was between change in ankle plantarflexor PROM and gastrocnemius MAS (R^2^ = 0.23), with decreases in spasticity after tSCS+SBLTT associated with increased PROM for all four participants (Figure 4B).

Self-selected gait speed increased throughout the study, with an average increase of 0.02 m/s after both SBLTT only and tSCS+SBLTT (Table 1). Changes in joint kinematics during gait varied between participants, but all participants had the greatest knee and hip extension post-tSCS+SBLTT or at tSCS+SBLTT follow-up (Figure 2 for example participant P03; Supplemental Figures for other participants). Three participants increased their hip extension throughout the gait cycle after tSCS+SBLTT compared to changes after SBLTT only. All participants had a greater increase in knee extension during stance after tSCS+SBLTT compared to after SBLTT only (Figure 3). Knee extension during swing increased after SBLTT only for three participants but increased after tSCS+SBLTT for all four participants. Three participants were more plantarflexed at the ankle during stance after tSCS+SBLTT, with their greatest ankle plantarflexion during push-off at the tSCS+SBLTT follow-up. The participant who initially walked in equinus-crouch became more dorsiflexed after tSCS+SBLTT (Figure 3; Supplemental Figure 2).

**Figure 2.**
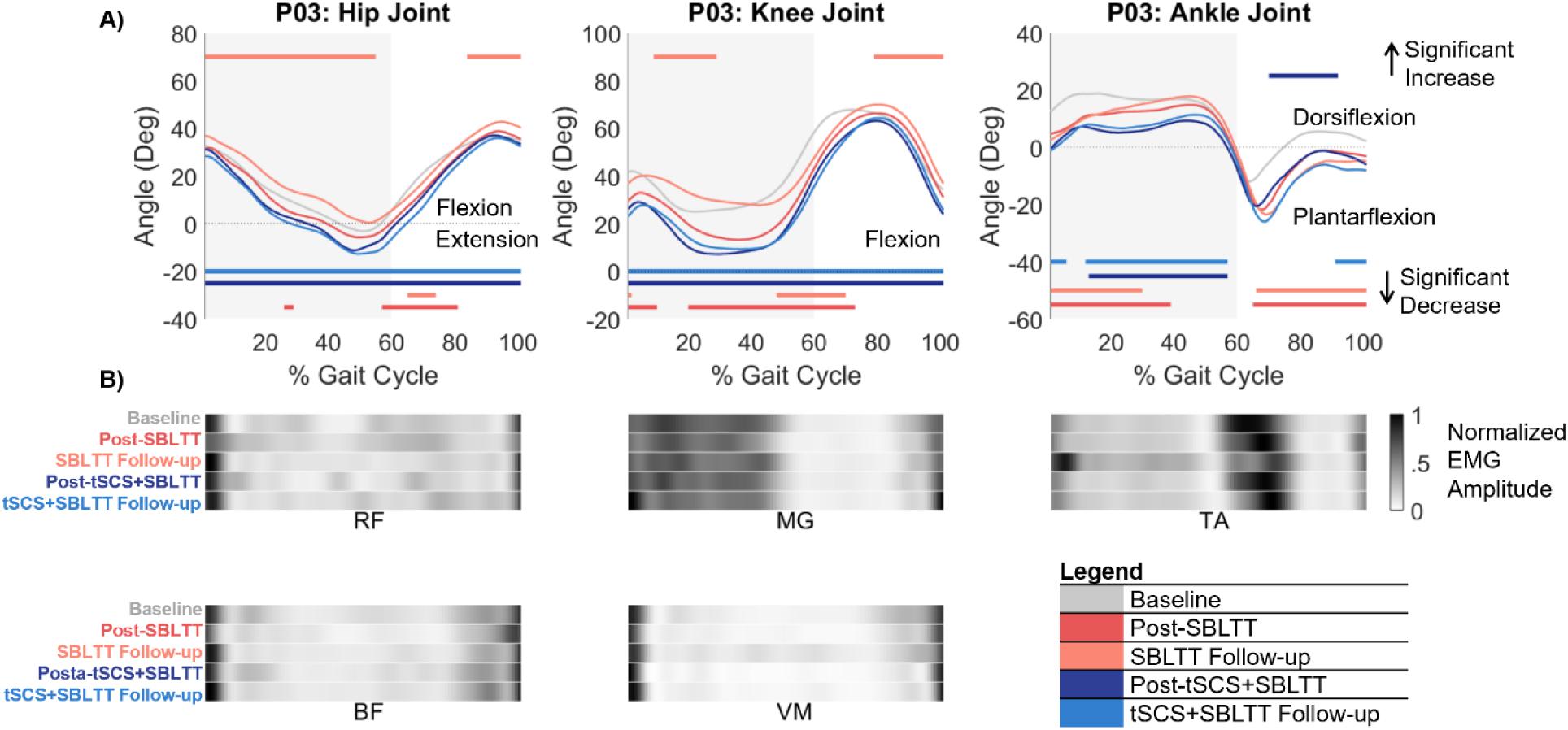
Example kinematics and muscles activity at each assessment timepoint for P03’s more-affected side. A) Sagittal-plane hip, knee, and ankle kinematics over the gait cycle. Horizontal colored lines indicate where there were significant changes in kinematics over each phase of the study based on statistical parametric mapping (p<0.05). Lines on top indicate locations of significant increases, while lines on the bottom indicate points of significant decreases. B) Normalized EMG amplitude during gait for the rectus femoris (RF), biceps femoris (BF), medial gastrocnemius (MG), vastus medialis (VM), and tibialis anterior (TA).

**Figure 3.**
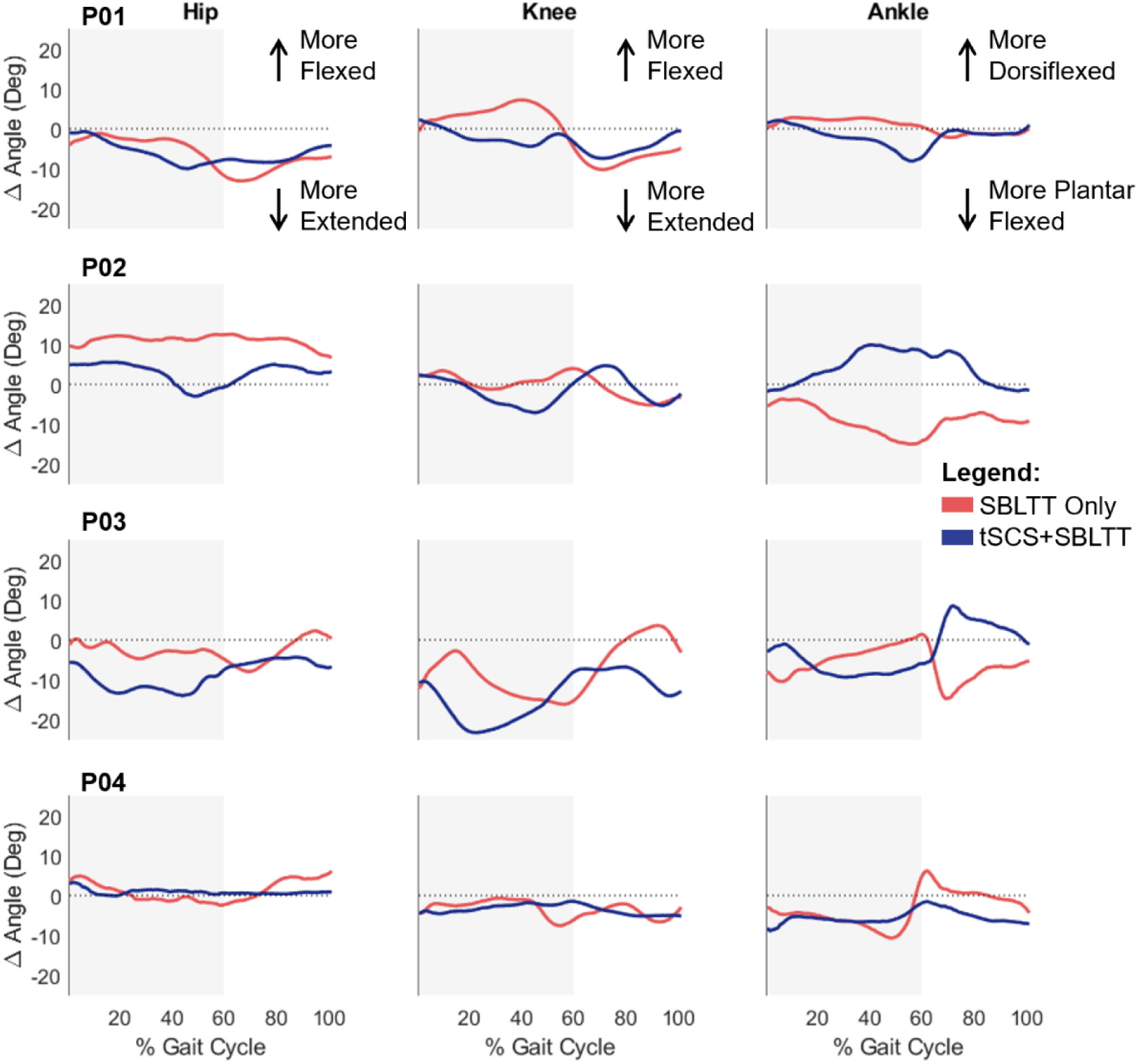
Change in the sagittal-plane joint kinematics over the gait cycle for the hip, knee, and ankle after SBLTT only (red) and tSCS+SBLTT (blue) for each participant. Positive values for the hip and knee indicate a more flexed position while negative values indicate a more extended position. A positive value for the ankle indicates a more dorsiflexed position and a negative value indicates a more plantarflexed position. The grey box on each figure indicates the stance phase of the gait cycle.

Despite increases in joint extension, there were minimal changes in the operating lengths of biarticular muscles at self-selected or fast gait speeds. We observed an average increase of 3.0% of the reference length and 0.13 nondimensional (n.d.) units in MTL and MTL rate, respectively, across muscles during SBLTT only and an average increase of 2.5% of the reference length and .0014 n.d. in MTL and MTL rate, respectively, during tSCS+SBLTT. Changes in MTLs were only weakly correlated with changes in spasticity at self-selected speeds (R^2^ < 0.18, Figure 4). At fast speeds, decreased gastrocnemius spasticity was associated with increased MTL (R^2^ = 0.26, Figure 4). There were no correlations between changes in spasticity and changes in MTL rate at either speed (R^2^ < 0.02).

**Figure 4.**
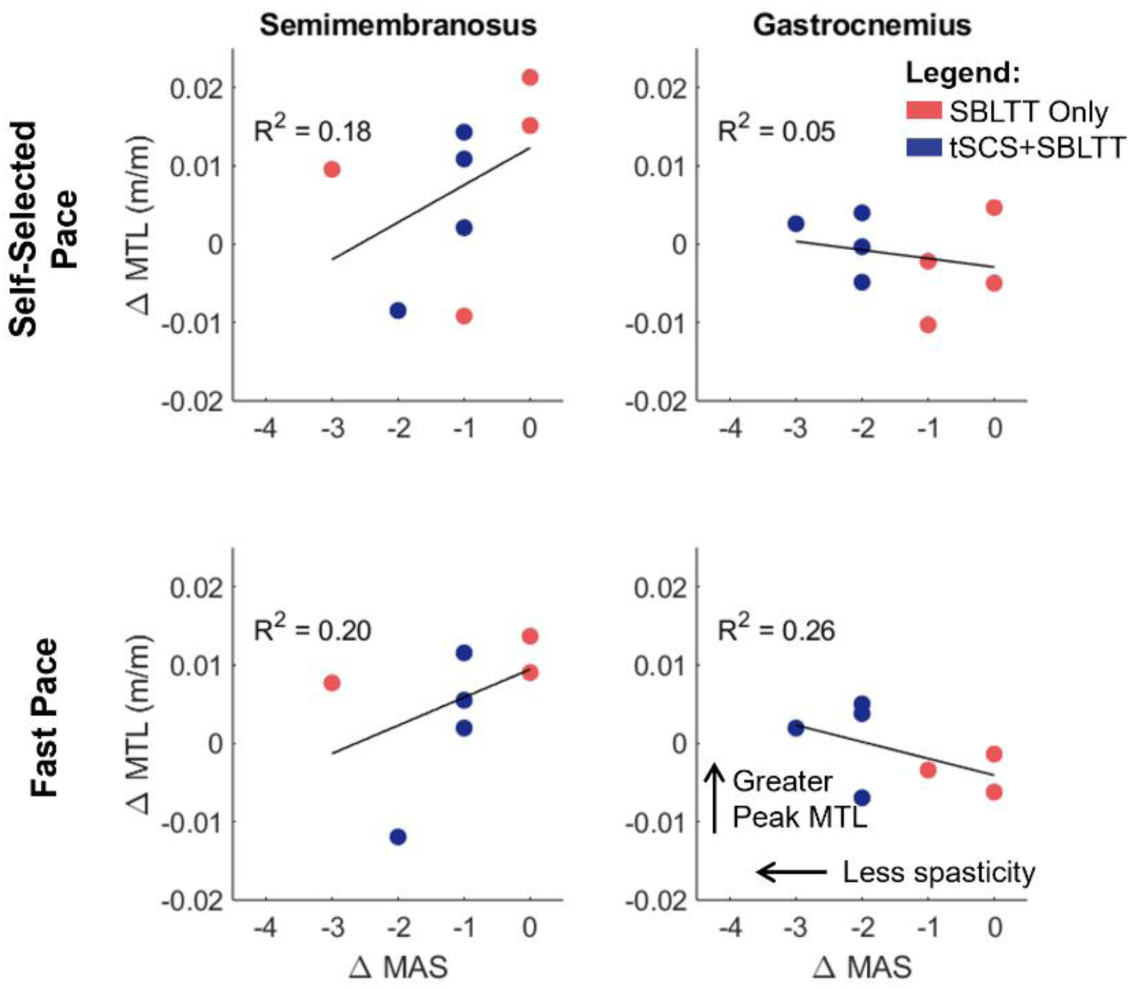
Linear regression comparing changes in operating musculotendon length (MTL) during self-selected and fast walking to changes in MAS for the semitendinosus and gastrocnemius muscles on the more affected side. MTLs are normalized to the muscle length when the participant is standing in an upright posture. Note: Fast pace is missing the ΔSBLTT data point for P01 because his fast speed was not recorded at baseline.

Changes in the timing and magnitude of muscle activations led to reduced co-contraction after tSCS+SBLTT compared to after SBLTT only (Table 1). The proximal muscles, vastus medialis and biceps femoris, had a 5% increase in CCI after SBLTT only and an 18% decrease in CCI after tSCS+SBLTT. The distal muscles, tibialis anterior and gastrocnemius, had a 0% change in CCI after SBLTT only and a 7% decrease in CCI after tSCS+SBLTT. Motor control during gait, as measured from muscle synergies, decreased slightly after SBLTT only and remained the same after tSCS+SBLTT (Table 1).

## Discussion

We hypothesized that reductions in spasticity after tSCS+SBLTT would correspond to changes in joint kinematics and muscle activity during gait for children with CP. The combination of tSCS+SBLTT led to more hip and knee extension and thereby more upright, less crouch position during walking. This new extended position may reduce demand on muscles, such as the reductions in antagonistic muscle co-contraction that we observed after tSCS+SBLTT, and potentially reduce muscle fatigue in CP [3,37]. Correlations between change in spasticity during each intervention and operating MTLs and PROM were generally weak, suggesting that there may have been other neuromuscular changes driving functional changes, such as muscle strength or selective motor control.

Like many children with CP, our participants exhibited characteristics of crouch gait at baseline with excessive knee flexion during stance and reduced hip extension, with one participant (P02) in equinus-crouch [37–39]. All participants exhibited their greatest knee extension during stance at the post-tSCS+SBLTT or tSCS+SBLTT follow-up timepoints. These results suggest that the addition of tSCS+SBLTT may support changes in muscle activity, such as reduced co-contraction and spasticity, that can enable greater knee extension and less fatiguing gait patterns [40,41] (Table 1). Future work should include assessments of muscle strength and selective motor control that may change with tSCS and contribute to the observed changes in gait pattern. Understanding the interacting effects of these impairments after intervention represents an important area for future research. Two participants also exhibited the greatest shifts in joint kinematics at the tSCS+SBLTT follow-up. While this may be due to continuing effects of the intervention, other environmental factors also likely played a role. For example, P01 started attending in-person school again after pandemic closures between the post-tSCS+SBLTT and the tSCS+SBLTT follow-up timepoint. His increase in daily walking activity may have further contributed to the positive changes in gait observed after the final non-intervention period [42].

Previous work has shown that spasticity may be one factor contributing to decreased musculotendon operating lengths and rates during gait for children with CP [17,27,28]. Van der Krogt et al. 2009 reported that hamstring and plantarflexor MTLs were 3-5% of the reference length shorter, and MTL rates of approximately 0.03 n.d. slower, for children with high spasticity and contracture compared to typically developing peers [27,28]. Another group reported that BoNT-A injections increased rectus femoris MTL by 5 mm and MTL rate by 0.2 m/s on the more spastic side [18]. We observed changes of similar magnitude, although only gastrocnemius MTL, not MTL rate, had a moderate correlation with decreased spasticity (Figure 4). It is possible that tSCS could have improved other neuromuscular impairments common in children with spastic CP, such as muscle weakness or selective motor control, which led to changes in MTL and MTL rates. Future work will need to include these outcomes simultaneously to better understand the possible mechanisms of tSCS.

Children with spasticity also have reductions in PROM that can be alleviated with spasticity treatments [43]. Carraro et al., 2014 reported that SDR significantly increased knee extension 13° and foot dorsiflexion 6° [19]. Choi et al., 2016 reported that BoNT-A injections significantly increased ankle range of motion 4° up to 4-months after injection [20]. We observed increases in range of motion and reductions in spasticity of comparable magnitudes. Correlations between PROM and MAS, however, were weak with the greatest correlation between PROM and MAS for the gastrocnemius (R^2^ = 0.23). This may be the strongest correlation because the positioning of the leg for PROM and MAS were identical, unlike other muscle groups. Measurement of PROM and MTL at the ankle can be influenced by foot morphology (i.e. midfoot break). To reduce these effects, we maintained the midfoot and subtalar at neutral for PROM and evaluated changes between sessions for individual participants in estimating MTLs at the ankle, since we expected minimal changes in foot morphology or properties over the course of training.

Clinically available treatments for spasticity may improve range of motion, but they have minimal or even detrimental effects on motor control [36]. Prior studies of muscle synergies reported a reduction in motor control complexity after SDR and BoNT-A injections (i.e., increase in tVAF_1_), suggesting that spasticity was masquerading as more complex muscle activity for children with CP. Our observation that tVAF_1_ stayed consistent during tSCS+SBLTT, despite significant reductions in spasticity, may suggest that tSCS+SBLTT has positive effects on motor control during gait relative to other approaches for spasticity management. As a result, selective motor control may be improved with tSCS+SBLTT, which should be considered in future studies.

As a preliminary study with four children with CP, this study has limited generalizability across the highly heterogeneous CP population. Rather, this study provides initial insight into how the addition of spinal stimulation and changes in spasticity may impact gait. We observed varied responses between individuals that likely depend on factors that should be considered in future studies such as age, baseline function (i.e. muscle weakness, MTL length, and selective motor control), stimulation intensity, and engagement during sessions. All analyses were performed on overground walking at self-selected or fast pace. There may be small effects of gait speed on outcomes like kinematics or MTLs when participants were instructed to walk at self-selected speed. However, average change in self-selected gait speed across timepoints was minimal (Table 1). Our participants had moderate spasticity comparable to children seeking spasticity care in clinical settings [19,20]. Several participants reached the lowest MAS scores, such that further potential effects on spasticity could not be quantified with MAS alone. Using instrumented spasticity tests may further elucidate the interplay between spasticity and gait biomechanics [44]. We recorded five muscles bilaterally for muscle activity and this did not include the gluteal muscles, but changes observed in hip kinematics could have been driven by changes in activity of the gluteal muscles.

The combination of tSCS and SBLTT resulted in reduction in spasticity, as well as changes in passive range of motion and joint kinematics during barefoot, overground walking in four children with CP. Changes in spasticity, however, do not explain all observations in the neuromechanical outcomes. More precise measures of spasticity combined with other assessments of neuromuscular impairments like weakness, muscle length, and impaired selective motor control are critical to guide effective treatment [45–47]. Future work should consider the neuromuscular factors leading to changes in joint movement, both passively and during walking, to further understand the mechanisms behind how tSCS can be optimized for children with CP. Non-invasive neuromodulation has the potential to decrease spasticity and improve walking in children with CP when combined with treadmill training.

## Data Availability

All data produced in the present study may be available upon reasonable request to the authors.

## Acknowledgements

The researchers would like to thank the families and research participants for their time and energy dedicated to this research. This work was supported by Seattle Children’s Hospital CP Research Pilot Study Fund 2020 Award, UW Rehabilitation Medicine Walter C. and Anita C. Stolov 2021 Research Fund, and NSF Graduate Research Fellowship Program Award DGE-1762114.

## Acknowledgement

The authors thank the children and their families for the time they dedicated to the research. We also thank Dr. Soshi Samejima, Rich Henderson, and Lauren Bachman for assisting with interventions and assessments, and Avocet Nagle-Christensen for assisting with data analysis.

