## Supplementary Figures for "Effects of spinal stimulation and short-burst treadmill training on gait biomechanics in children with cerebral palsy"

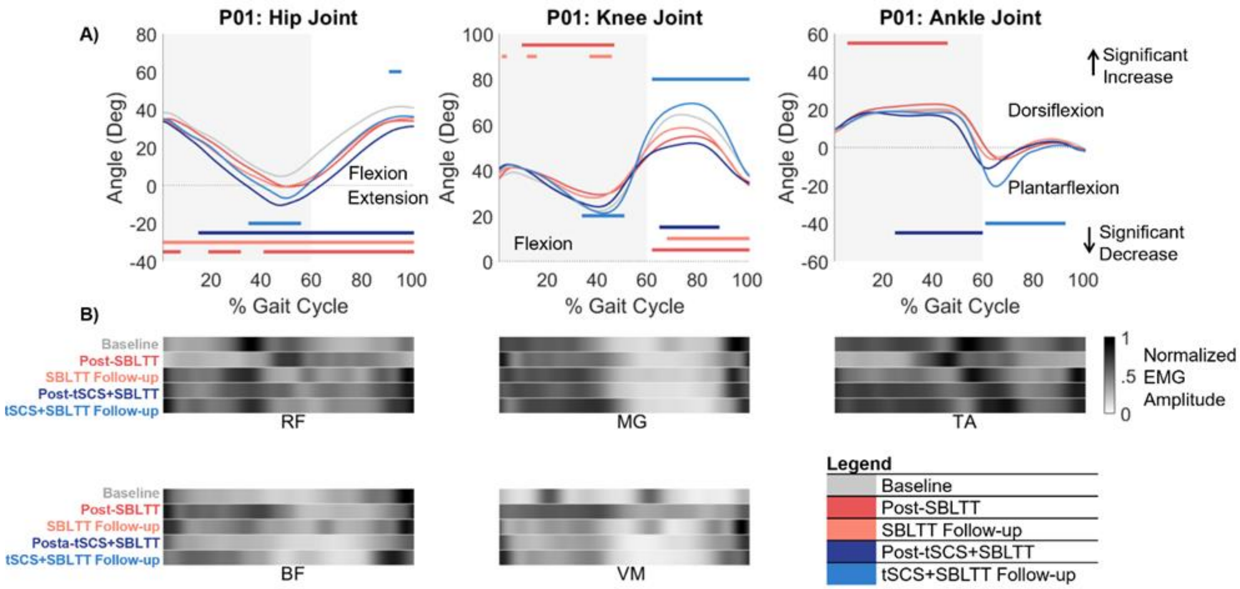


**Supplemental Figure 1.** P01's more-affected side. A) Sagittal-plane hip, knee, and ankle kinematics over the gait cycle. Horizontal colored lines indicate where there were significant changes in kinematics over each phase of the study based on statistical parametric mapping ($p<0.05$). Lines on top indicate locations of significant increases, while lines on the bottom indicate points of significant decreases. B) Normalized EMG amplitude during gait for the lower extremity muscles.

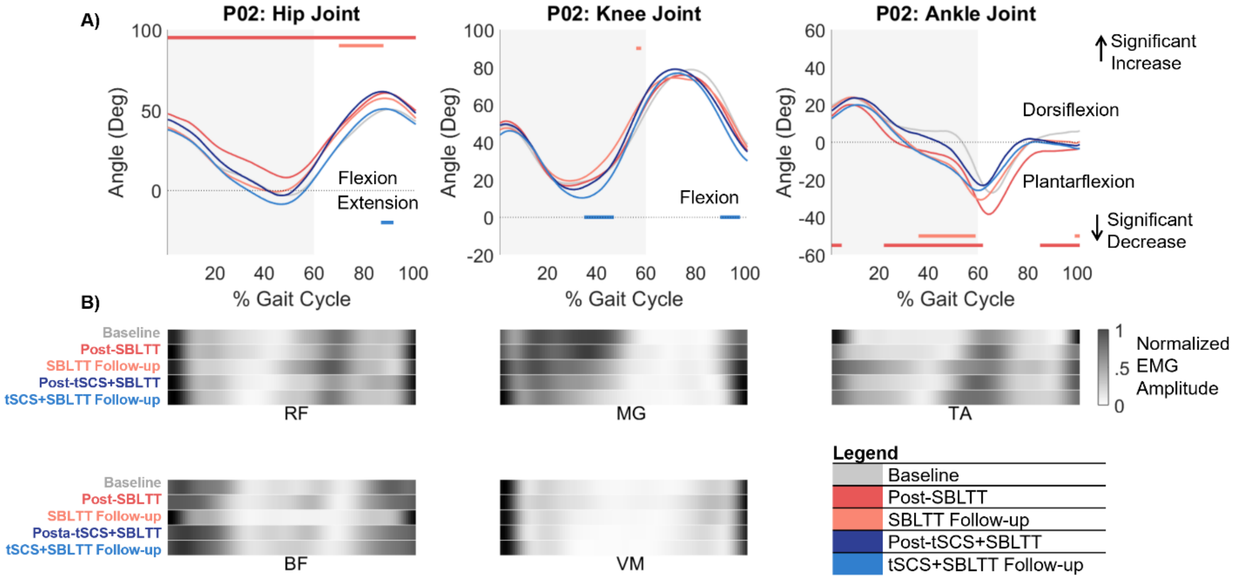


**Supplemental Figure 2.** P02's more-affected side. A) Sagittal-plane hip, knee, and ankle kinematics over the gait cycle. Horizontal colored lines indicate where there were significant changes in kinematics over each phase of the study based on statistical parametric mapping (p<0.05). Lines on top indicate locations of significant increases, while lines on the bottom indicate points of significant decreases. B) Normalized EMG amplitude during gait for the lower extremity muscles.

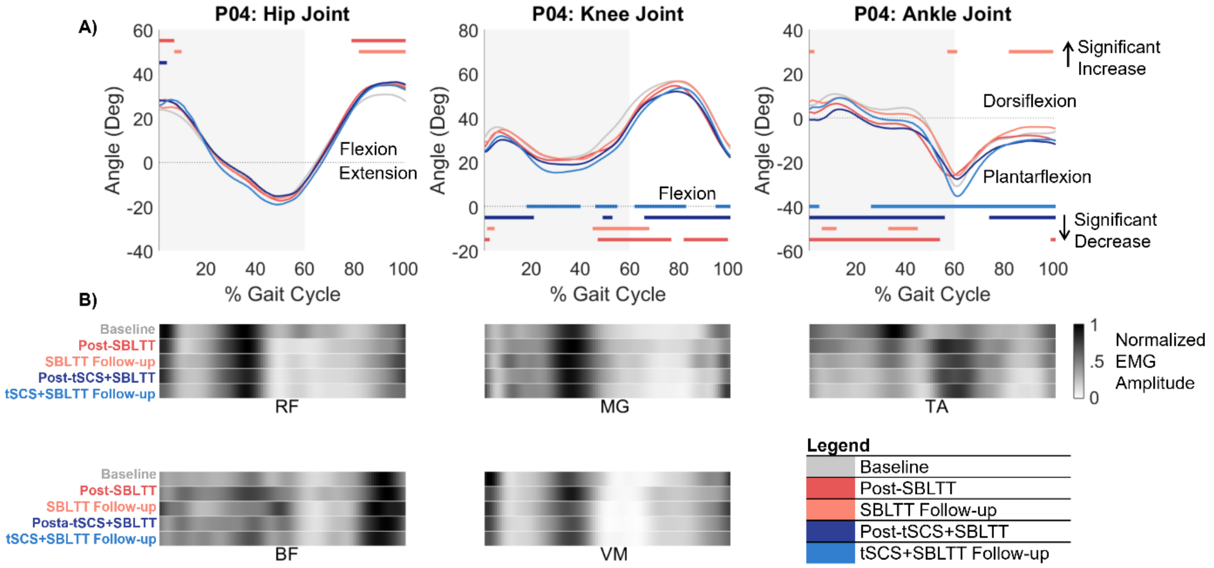


**Supplemental Figure 3.** P04's more-affected side. A) Sagittal-plane hip, knee, and ankle kinematics over the gait cycle. Horizontal colored lines indicate where there were significant changes in kinematics over each phase of the study based on statistical parametric mapping (p<0.05). Lines on top indicate locations of significant increases, while lines on the bottom indicate points of significant decreases. B) Normalized EMG amplitude during gait for the lower extremity muscles.

**Supplemental Table 1.** Spasticity, PROM, and gait measures at each time point for P01.

| | | Baseline | Post-SBLTT | SBLTT Follow-up | Post-tSCS+SBLTT | tSCS+SBLTT Follow-up |
|---|---|---|---|---|---|---|
| **MAS** | Quadriceps | 3 | 2 | 2 | 0 | 0 |
| | Hamstrings | 2 | 1 | 2 | 0 | 1 |
| | Gastrocnemius | 3 | 2 | 3 | 1 | 1 |
| | Soleus | 4 | 3 | 4 | 2 | 2 |
| **PROM (degrees)** | Hip Flexion | 37 | 43 | 48 | 45 | 38 |
| | Knee Extension | 165 | 154 | 127 | 162 | 159 |
| | Knee Flexion | 121 | 126 | 110 | 110 | 125 |
| | Ankle Dorsiflexion | 75 | 76 | 60 | 60 | 76 |
| | Ankle Plantarflexion | 120 | 140 | 145 | 155 | 158 |
| **Gait Speed (n.d.)** | Self-selected Pace | 0.31 | 0.31 | 0.37 | 0.41 | 0.39 |
| | Fast Pace | * | 0.50 | 0.54 | 0.44 | 0.51 |
| **Integrated EMG During Stance [% of gait cycle used]** | RF [0-60] | 31 | 24 | 33 | 33 | 33 |
| | VM [0-60] | * | 43 | 24 | 21 | 26 |
| | BF [0-60] | 19 | 37 | 29 | 18 | 27 |
| | MG [30-50] | 8 | 11 | 11 | 10 | 15 |
| | TA [6-60] | 37 | 26 | 29 | 35 | 36 |
| **CCI (%)** | Proximal Muscles | * | 86 | 81 | 81 | 75 |
| | Distal Muscles | 75 | 64 | 74 | 79 | 85 |
| **Motor Control** | $tVAF_1$ | * | 0.93 | 0.92 | 0.92 | 0.89 |

*P01 is not included in the baseline fast walking speed or EMG of the VM, which also resulted in no calculation of proximal CCI or $tVAF_1$.

**Supplemental Table 2.** Spasticity, PROM, and gait measures at each time point for P02.

| | | Baseline | Post-SBLTT | SBLTT Follow-up | Post-tSCS+SBLTT | tSCS+SBLTT Follow-up |
|---|---|---|---|---|---|---|
| **MAS** | Quadriceps | 1 | 0 | 1 | 0 | 0 |
| | Hamstrings | 1 | 1 | 1 | 0 | 1 |
| | Gastrocnemius | 4 | 3 | 4 | 2 | 2 |
| | Soleus | 4 | 4 | 4 | 2 | 2 |
| **PROM (degrees)** | Hip Flexion | 42 | 42 | 47 | 64 | 42 |
| | Knee Extension | 104 | 154 | 166 | 174 | 167 |
| | Knee Flexion | 153 | 125 | 133 | 150 | 130 |
| | Ankle Dorsiflexion | 50 | 77 | 60 | 95 | 80 |
| | Ankle Plantarflexion | 146 | 168 | 162 | 160 | 174 |
| **Gait Speed (n.d.)** | Self-selected Pace | 0.50 | 0.58 | 0.59 | 0.58 | 0.60 |
| | Fast Pace | 0.53 | 0.57 | 0.70 | 0.64 | 0.75 |
| **Integrated EMG During Stance [% of gait cycle used]** | RF [0-60] | 17 | 18 | 17 | 16 | 17.5 |
| | VM [0-60] | 13 | 14 | 12 | 11 | 12 |
| | BF [0-60] | 23 | 22 | 15 | 27 | 26 |
| | MG [30-50] | 14 | 13 | 8 | 6 | 4 |
| | TA [6-60] | 10 | 13 | 25 | 17 | 23 |
| **CCI (%)** | Proximal Muscles | 52 | 53 | 68 | 37 | 36 |
| | Distal Muscles | 34 | 44 | 62 | 52 | 58 |
| **Motor Control** | $tVAF_1$ | 0.72 | 0.76 | 0.80 | 0.81 | 0.81 |

**Supplemental Table 3.** Spasticity, PROM, and gait measures at each time point for P03.

| | | Baseline | Post-SBLTT | SBLTT Follow-up | Post-tSCS+SBLTT | tSCS+SBLTT Follow-up |
|---|---|---|---|---|---|---|
| **MAS** | Quadriceps | 1 | 0 | 0 | 0 | 0 |
| | Hamstrings | 3 | 0 | 1 | 0 | 1 |
| | Gastrocnemius | 4 | 4 | 4 | 1 | 1 |
| | Soleus | 3 | 3 | 3 | 1 | 1 |
| **PROM (degrees)** | Hip Flexion | 30 | 45 | 35 | 45 | 35 |
| | Knee Extension | 156 | 155 | 152 | 155 | 165 |
| | Knee Flexion | 125 | 135 | 130 | 130 | 125 |
| | Ankle Dorsiflexion | 72 | 70 | 80 | 90 | 85 |
| | Ankle Plantarflexion | 155 | 160 | 130 | 152 | 150 |
| **Gait Speed (n.d.)** | Self-selected Pace | 0.50 | 0.40 | 0.36 | 0.43 | 0.49 |
| | Fast Pace | 0.60 | 0.59 | 0.60 | 0.62 | 0.64 |
| **Integrated EMG During Stance [% of gait cycle used]** | RF [0-60] | 17 | 20 | 15 | 16 | 17 |
| | VM [0-60] | 11 | 8 | 13 | 7 | 10 |
| | BF [0-60] | 16 | 10 | 13 | 15 | 15 |
| | MG [30-50] | 13 | 9 | 12 | 12 | 11 |
| | TA [6-60] | 13 | 12 | 17 | 10 | 10 |
| **CCI (%)** | Proximal Muscles | 68 | 66 | 81 | 44 | 59 |
| | Distal Muscles | 42 | 48 | 62 | 43 | 51 |
| **Motor Control** | $tVAF_1$ | 0.73 | 0.71 | 0.74 | 0.74 | 0.77 |

**Supplemental Table 4.** Spasticity, PROM, and gait measures at each time point for P04.

| | | Baseline | Post-SBLTT | SBLTT Follow-up | Post-tSCS+SBLTT | tSCS+SBLTT Follow-up |
|---|---|---|---|---|---|---|
| **MAS** | Quadriceps | 0 | 0 | 0 | 0 | 0 |
| | Hamstrings | 1 | 1 | 1 | 0 | 0 |
| | Gastrocnemius | 3 | 3 | 3 | 1 | 1 |
| | Soleus | 3 | 3 | 3 | 1 | 1 |
| **PROM (degrees)** | Hip Flexion | 35 | 45 | 30 | 42 | 38 |
| | Knee Extension | 145 | 155 | 150 | 164 | 165 |
| | Knee Flexion | 130 | 130 | 120 | 138 | 140 |
| | Ankle Dorsiflexion | 68 | 55 | 65 | 78 | 65 |
| | Ankle Plantarflexion | 150 | 150 | 160 | 155 | 145 |
| **Gait Speed (n.d.)** | Self-selected Pace | 0.52 | 0.60 | 0.55 | 0.53 | 0.57 |
| | Fast Pace | 0.75 | 0.90 | 0.83 | 0.88 | 0.92 |
| **Integrated EMG During Stance [% of gait cycle used]** | RF [0-60] | 31 | 26 | 26 | 25 | 23 |
| | VM [0-60] | 25 | 26 | 22 | 21 | 22 |
| | BF [0-60] | 20 | 31 | 26 | 26 | 26 |
| | MG [30-50] | 12 | 15 | 15 | 14 | 14 |
| | TA [6-60] | 31 | 30 | 23 | 19 | 17 |
| **CCI (%)** | Proximal Muscles | 62 | 64 | 63 | 60 | 63 |
| | Distal Muscles | 77 | 74 | 71 | 64 | 60 |
| **Motor Control** | $tVAF_1$ | 0.87 | 0.86 | 0.84 | 0.80 | 0.81 |
